# Modelling serological multiplex bead assays responses: A case study from Malaysia

**DOI:** 10.64898/2025.12.11.25342063

**Authors:** Chiara Romano, Jonas Wallin, Timothy William, Chris Drakeley, Branda Francesco, Massimo Ciccozzi, Emanuele Giorgi

## Abstract

**Background:** Multiplex bead assays (MBAs) provide quantitative measurements of many analytes from small sample volumes, reducing cost and processing time compared with traditional immunoassays. These advantages have made MBAs valuable for studying diverse diseases, particularly in low-resource settings. However, most analytical approaches focus on individual diseases, while integrated surveillance platforms would benefit from methods that jointly analyse the full range of pathogens included in multiplex assays.

**Methods:** We applied factor analysis combined with a Dirichlet process mixture model to identify sub-populations based on MBA responses and assess whether these groups show spatial patterns or share socioeconomic characteristics and disease exposures. Data were drawn from four districts in northern Sabah, Malaysia, and included antibody responses for several neglected tropical diseases (NTDs): strongyloidiasis, lymphatic filariasis, giardiasis, toxoplasmosis, trachoma, and yaws.

**Results:** The model identified four distinct sub-populations. Three of these were spatially distributed and included people with similar socioeconomic profiles. These shared characteristics may help explain the antibody patterns observed within each group, offering a comprehensive characterization of each sub-population.

**Conclusion:** The presented analytical workflow, combining factor analysis, Dirichlet process mixture modelling, and spatial analysis, offers a replicable approach for identifying multi-disease exposure clusters that could inform integrated, targeted interventions.

**Author summary:** Serological antibody levels, i.e. the amount of pathogen-specific antibodies in the blood, provide a cost-effective way to measure past exposure to infectious diseases. This study analyzed antibody measurements of several neglected tropical diseases of a sampled population in northern Sabah, Malaysia. Our goal was to identify sub-populations that share similar disease exposure profiles and socioeconomic characteristics. To find these groups, we used a novel unsupervised learning approach, which allows the data to reveal patterns naturally without prior assumptions. The method identified four distinct sub-populations. Three were clearly defined: two were primarily driven by antibodies to a single pathogen (indicating one dominant infection), and one showed low antibody levels across all pathogens, indicating generally low exposure. These sub-groups were also linked to specific geographic locations and socioeconomic factors, suggesting that where people live and their specific living conditions help explain different infection patterns. This approach allows relationships between pathogens and exposure risks to emerge directly from the data. It can help integrated surveillance systems identify where and in whom multiple infections occur, enabling public health teams to better target testing.

## Introduction

Serological antibody levels serve as sensitive indicators of pathogen exposure [1], and their measurement has become increasingly important in studying and managing a wide range of diseases [2]. Multiplex bead assays (MBA) provide quantitative measurement of a large number of analytes using small sample volumes, thereby reducing both cost and processing time compared to traditional immunoassays [2, 3]. When integrated into existing surveillance platforms, MBAs can help the global community to identify public health gaps more efficiently [1], support the development of targeted interventions, identify potential risk factors and determine populations eligible for vaccination [3]. Integrated serological surveillance using MBAs has proven particularly effective in low– and middle-income countries [3] because of their rapid, precise, and cost-effective measurement of current and past pathogen exposure [2, 4].

Despite these advantages, the analysis of MBA data poses several challenges. The large number of analytes introduces both data-quality issues due to extensive pre-processing and normalization [5, 6], and difficulties in interpretation, as understanding patterns across multiple diseases demands interdisciplinary expertise. Most studies to date have focused on analyzing MBA data one disease at a time [3], leaving a gap in our understanding of how multiplex data can reveal broader population-level patterns of exposure and co-endemicity. Yet, integrated surveillance platforms would greatly benefit from analytical frameworks capable of identifying population subgroups or geographical areas characterized by multiple co-occurring diseases [1].

In this work, we employ multivariate clustering methods to address this analytical gap, leveraging the information contained across multiple diseases in the multiplex data. Specifically, our goal is to determine whether the quantitative signals produced by multiplex assays can be used to identify groups of individuals (*clusters* or *sub-populations*) who share similar serological profiles, which we can interpret as distinct health profiles, and to characterise these profiles in terms of demographic, socio-economic, and spatial variables. This perspective shifts the focus from single-disease analyses toward a data-driven understanding of population-level health structure.

A number of studies have sought to extract epidemiologically relevant information from multiplex data using a variety of analytical approaches. Early work by Arnold et al. fitted age-dependent antibody curves to multiplex responses to estimate changes in transmission intensity over time [1], while Macalinao et al. combined statistical and machine-learning models to distinguish recent from historical exposure in low-transmission settings [7]. Clarke et al. used mixed-effects models to normalise MBA data for technical variance arising from plate, batch, or assay-run effects [5], and Matson et al. developed a Shiny-based platform to facilitate data quality control and visual inspection [6]. More recently, researchers have begun to explore the use of multivariate methods to summarise and interpret multiplex responses across pathogens. Schultz et al. applied Principal Component Analysis (PCA) to derive composite indicators of cumulative exposure [8]; Carcelen et al. used multiple logistic regression to quantify cross-pathogen seropositivity and examine the influence of demographic and spatial factors [9]; and Fornace et al. integrated multiplex data with geostatistical models to map exposure to multiple neglected tropical diseases (NTDs) in Northern Ghana [10]. However, while these approaches make use of the full range of diseases included in MBA data, none employ analytical methods that begin at the population level to uncover latent groups of individuals who share similar serological profiles across multiple pathogens.

Building on this body of work, our analysis combines factor analysis and clustering through a Dirichlet process mixture model to identify sub-populations and their spatial distribution. This framework integrates the strengths of previous approaches (e.g. dimensionality reduction [8], multi-pathogen analysis [9], and spatial mapping of exposure [10]) to uncover latent health profiles within the population. It thus offers a unified, data-driven approach to examine how multiple exposures interact, and how these interactions relate to socio-economic characteristics and geography.

We apply this analytical framework to multiplex data on NTDs collected in Malaysia, including strongyloidiasis, lymphatic filariasis (LF), giardiasis, toxoplasmosis, trachoma and yaws. Over the past decade, Malaysia’s mass-drug administrations, sanitation improvements, and public-health initiatives have reduced these NTDs, yet they remain endemic particularly in isolated, resource-limited, and aboriginal communities in Sabah [11, 12], posing a continued challenge to the elimination and eradication goals set by the WHO NTD Roadmap 2021–2030 [13]. Given that the transmission of many of these pathogens geographically overlaps, often resulting in co-infections [14], an integrated, multi-disease analysis offers new perspectives beyond those achievable through single-disease approaches. The scope of this work is therefore to present a reproducible workflow for analysing multiplex antibody data that enables the identification and characterisation of health profiles within populations. By uncovering these clusters, we aim to enhance the interpretability of MBA data, unravel exposure patterns at the population-level, and ultimately support the development of evidence-based strategies for integrated surveillance.

## Materials and methods

### Data description

The study utilized data from a previously conducted environmentally stratified, population-based, cross-sectional serological survey carried out in four districts of northern Sabah, Malaysia (see Figure 1) between September 17 and December 12, 2015, as described by Fornace et al. [15]. A non-self-weighting two-stage sampling design targeted 2,650 households across 919 villages (average population 90 individuals, 36 households) stratified according to the proportion of forest cover in 2014 within a 2 km radius of the village centroid, excluding urban areas. All individuals residing in sampled households were asked to participate (ages 3 months-105 years). The geographical location of households was recorded in the WGS84/UTM zone 50N projected coordinate reference system (EPSG:32650).

**Fig 1.**
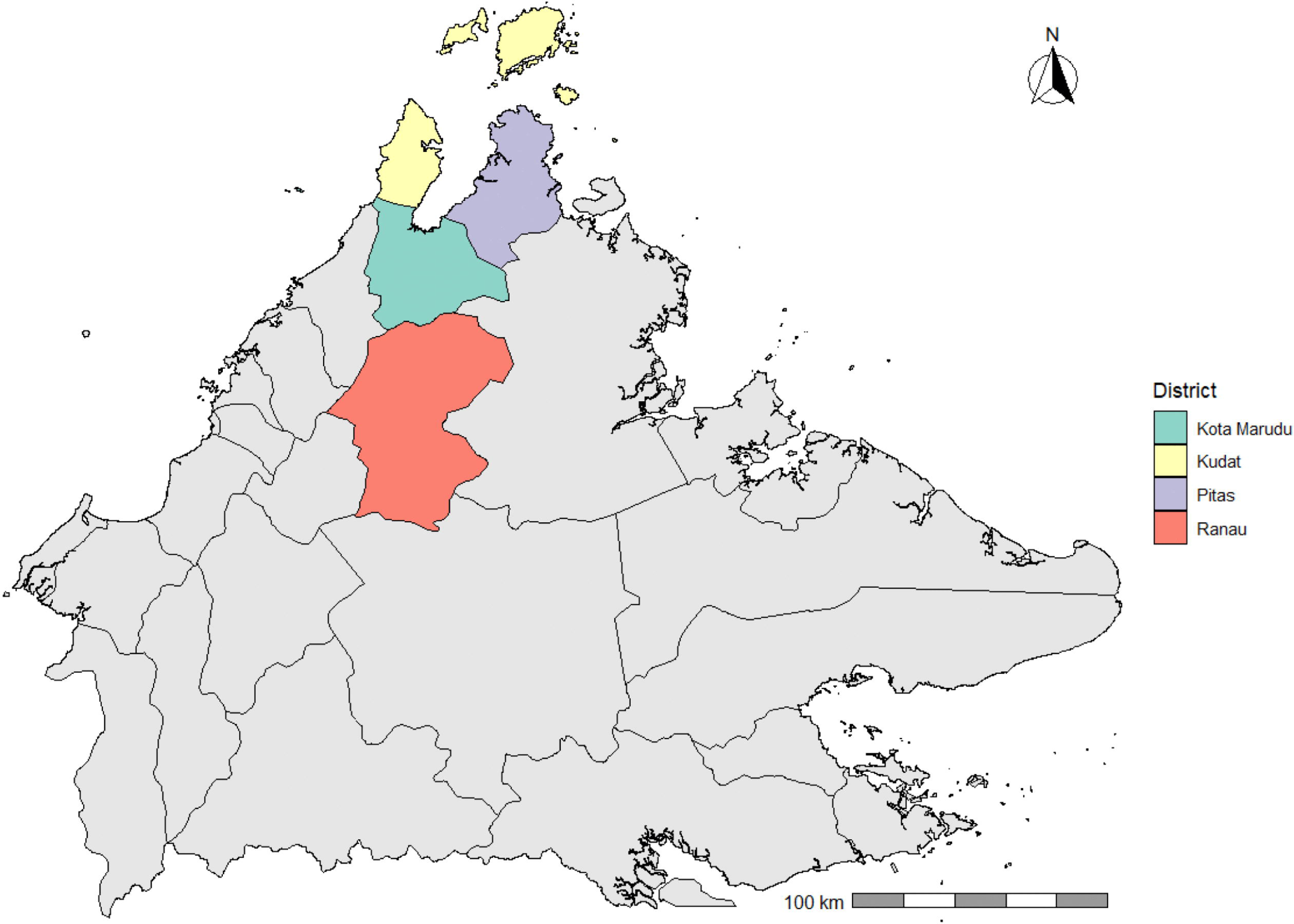
Study area. Study area in northern Sabah, Malaysia.

The dataset, resulting in a total of 6,168 observations from 1,905 unique household locations, included individual-level variables such as age, sex, and occupation as well as household-level variables, including population density, income, and vegetation stratum. Descriptive statistics for these variables are provided in Supplementary Material Table S1. Serological multiplex bead assays measured the IgG antibody responses to twelve antigens from six pathogens: lymphatic filariasis (Bm33, Bm14, BmR1, Wb123), strongyloidiasis (NIE), toxoplasmosis (SAG2A), yaws (Rp17, TmpA), trachoma (Pgp3, Ct694), and giardiasis (VSP3, VSP5). To quantify the IgG responses, Median fluorescence intensity minus background (MFI-bg) was measured using a Luminex MAGPIX bioanalyzer and xPONENT software (version 4.2), following protocols detailed by Chan et al. [14].

To provide context for the integrated analysis, the pathogens included in the MBA panel represent a mix of helminth, protozoan, and bacterial infections: Table 1 provides a brief description of the main symptoms and primary modes of transmission, the specific antigens and their epidemiological interpretation associated with each disease. However, it should be noted that these antigen-specific interpretations reflect general patterns observed at the population level; individual antibody responses, and their decay kinetics over time, may vary due to host-specific immunological factors.

**Table 1.**
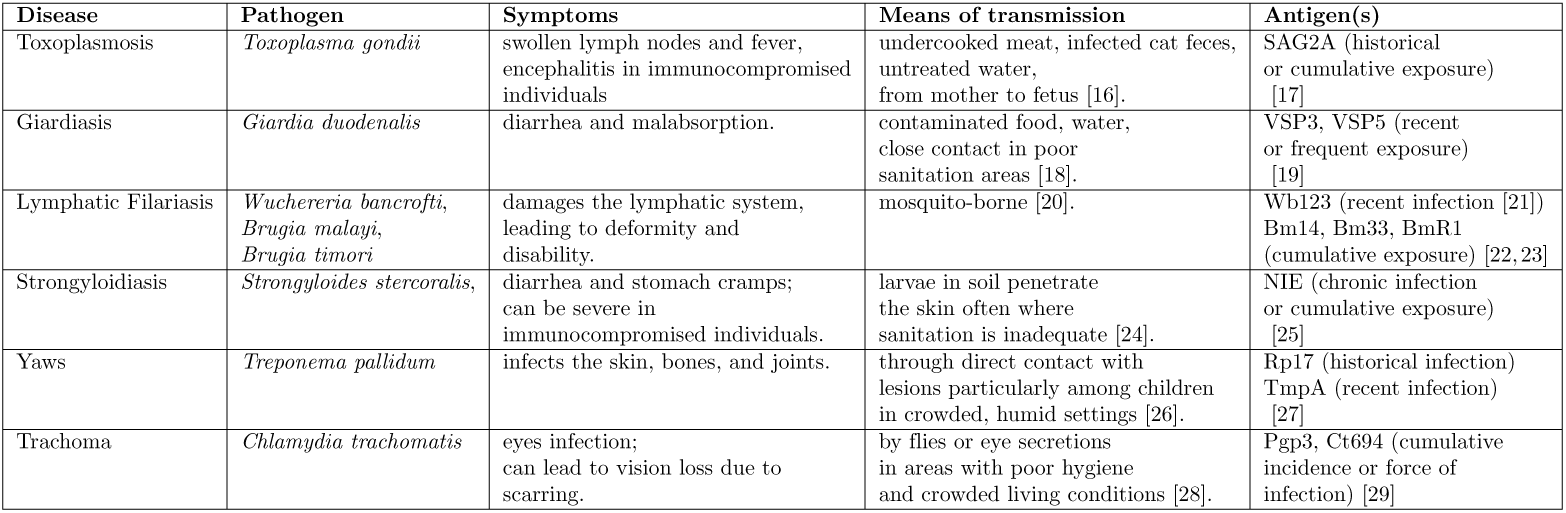
Short description of the pathogens included in the analysis.

| Disease | Pathogen | Symptoms | Means of transmission | Antigen(s) |
| --- | --- | --- | --- | --- |
| Toxoplasmosis | *Toxoplasma gondii* | swollen lymph nodes and fever, encephalitis in immunocompromised individuals | undercooked meat, infected cat feces, untreated water, from mother to fetus [16]. | SAG2A (historical or cumulative exposure) [17] |
| Giardiasis | *Giardia duodenalis* | diarrhea and malabsorption. | contaminated food, water, close contact in poor sanitation areas [18]. | VSP3, VSP5 (recent or frequent exposure) [19] |
| Lymphatic Filariasis | *Wuchereria bancrofti*, *Brugia malayi*, *Brugia timori* | damages the lymphatic system, leading to deformity and disability. | mosquito-borne [20]. | Wb123 (recent infection [21])<br>Bm14, Bm33, BmR1 (cumulative exposure) [22, 23] |
| Strongyloidiasis | *Strongyloides stercoralis*, | diarrhea and stomach cramps; can be severe in immunocompromised individuals. | larvae in soil penetrate the skin often where sanitation is inadequate [24]. | NIE (chronic infection or cumulative exposure) [25] |
| Yaws | *Treponema pallidum* | infects the skin, bones, and joints. | through direct contact with lesions particularly among children in crowded, humid settings [26]. | Rp17 (historical infection)<br>TmpA (recent infection) [27] |
| Trachoma | *Chlamydia trachomatis* | eyes infection; can lead to vision loss due to scarring. | by flies or eye secretions in areas with poor hygiene and crowded living conditions [28]. | Pgp3, Ct694 (cumulative incidence or force of infection) [29] |

### Statistical Analysis

The methodology employed in this work consists of a three-step analysis: (i) first, we performed dimensionality reduction on the antigens’ quantitative antibody levels; (ii) second, we clustered the resulting reduced matrix to identify sub-populations (clusters); and (iii) finally, we estimated the spatial probability of belonging to each cluster. Before carrying out the steps above, all the log-transformed MFI-bg values are standardized.

All analyses were conducted using R Statistical Software (v4.4.3; R Core Team 2021) [30].

#### Dimensionality reduction

Dimensionality reduction was applied to generate a representative indicator for each pathogen. This approach allowed us to focus on the relationships between diseases rather than individual antigens and to reduce the collinearity between antigens (see the correlation matrix in Figure S1 in the Additional file 1), which can influence both the shape and number of identified clusters [31].

We performed dimensionality reduction using factor analysis on the scaled log-transformed MFI-bg values for each group of antigens associated with the same disease. Specifically, we analyzed four groups: the four antigens Bm33, Bm14, BmR1, and Wb123 for lymphatic filariasis; the two antigens Rp17 and TmpA for yaws; the two antigens Pgp3 and Ct694 for trachoma; and the two antigens VSP3 and VSP5 for giardiasis.

To determine the number of factors for each antigen group, we used both a scree plot examination and the Kaiser Criterion (eigenvalues greater than one) [32]. Both criteria suggested a single factor for all four antigen groups, which we will later refer to by the name of the associated disease. Since rotation techniques only redistribute variance across multiple factors, no rotation was applied to the one-factor solutions.

The result of this first step was a six-dimensional data matrix, with each column representing a disease.

#### Cluster analysis

We applied a Dirichlet Process Mixture Model (DPMM) for cluster analysis on the diseases six-dimensional data matrix. DPMM is a bayesian nonparametric method and a Model-based clustering technique, meaning that each cluster corresponds to a mixture component described by a probability distribution with unknown parameters. In this work the mixture components are assumed to be distributed as Gaussian. Unlike traditional methods, such as k-means, which require the number of clusters to be fixed in advance, DPMM automatically infers the number of clusters *k* based on the data. Moreover, while traditional approaches typically yield “hard” assignments of data points to clusters, DPMM provides probabilistic cluster memberships, capturing the inherent uncertainty in the data [33]. Intuitively, the model can be understood as a two-step, sequential process. (1) Individuals are allocated to clusters one at a time: each new individual either joins one of the clusters already formed by previous individuals, with a probability proportional to that cluster’s current size, or starts a brand-new cluster, with a probability governed by a concentration parameter *α* (larger values of *α* make the creation of new clusters more likely). (2) Once cluster membership is determined, each individual’s six-dimensional disease profile is assumed to arise from a Gaussian distribution centred on the mean and covariance specific to that cluster. We have *n* observations **y**_1_*,..,* **y***_n_* where 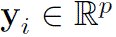 and *p* = 6, the number of diseases in our example. Let 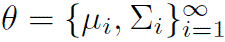 be the mixture components’ parameters.

Briefly the DPMM could be written as:

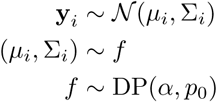

where the first states that each individual’s disease profile *y_i_* is generated from a Gaussian distribution with mean *µ_i_* and covariance Σ*_i_*, both specific to the cluster the individual belongs to; the second states that these cluster-specific parameters are themselves drawn from an (unknown) distribution *f*; and the third specifies that *f* is itself random, drawn from a Dirichlet Process (DP) with concentration parameter *α* and base distribution *p*_0_. It is this DP prior placed on the cluster-specific parameters *θ* that accounts for a theoretically infinite number of mixture components. In real-life applications, however, the number of mixture components (i.e. clusters) cannot exceed the number of observations *n* so that 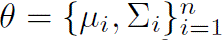. In this work, we assumed homoscedastic and isotropic covariances: 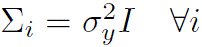 and 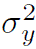 is a fixed parameter.

Our R implementation of the DPMM and the following sensitivity analysis, was adapted from the methodological framework proposed by Li et al. [33]. To select the DP concentration parameter *α* and the within-cluster variance 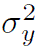, we performed a sensitivity analysis. We estimated a DPMM model for all combinations of *α* = (0.01, 1.00, 2.00, 3.00) and 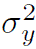 = (0.50, 1.00, 1.5, 3.00) to examine their effect on clustering. Large *α* values encourage more clusters, while larger 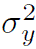 values lead to fewer clusters. The chosen *α* values cover a range of small and large values, with *α* = 1 being neutral in the creation of new clusters. The 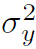 values were selected based on a visual examination of the scatterplots between each pair of variables (see Figure S2 in the Additional file 1), capturing a range of small and large variability.

To determine the best combination of *α* and 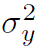, we compared the different DPMM models using four internal cluster validity measures: Silhouette, Dunn, Davies–Bouldin (DB), and Calinski–Harabasz (CH) indices. We also considered the algorithm’s variability in selecting the final number of clusters, measured by the standard deviation of the number of clusters over the iterations, sd(k) (fifth column in Table 2). Higher values for Silhouette, Dunn, and CH indicate better clustering, while lower values for DB and *sd*(*k*) suggest a better classification.

**Table 2.** Sensitivity analysis to choose the DPMM’ parameters *α* and 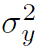. Optimal values for each measure is shown in bold. Order given by the Silhouette score.

| $\alpha$ | $\sigma_y^2$ | $k$ | $mean(k)$ | $sd(k)$ | Silhouette | Dunn | DB | CH |
| --- | --- | --- | --- | --- | --- | --- | --- | --- |
| 0.01 | 1.5 | 2 | 2.342 | **0.479** | 0.526 | 0.082 | 0.740 | 166.5 |
| 3.00 | 1.5 | 4 | 6.125 | 1.439 | **0.382** | **0.046** | **0.914** | 283.8 |
| 2.00 | 1.5 | 4 | 5.426 | 1.193 | 0.382 | **0.046** | 0.917 | 283.9 |
| 1.00 | 1.5 | 4 | 4.620 | 0.844 | 0.381 | **0.046** | 0.917 | 286.3 |
| 0.01 | 1.0 | 5 | 4.454 | 0.550 | 0.232 | 0.030 | 1.404 | **669.2** |
| 2.00 | 1.0 | 6 | 7.684 | 1.296 | 0.213 | 0.020 | 1.34 | 611.8 |
| 1.00 | 1.0 | 6 | 7.058 | 0.986 | 0.209 | 0.025 | 1.341 | 631.7 |
| 0.01 | 0.5 | 13 | 11.938 | 1.597 | 0.158 | 0.024 | 1.416 | 758.9 |
| 1.00 | 0.5 | 25 | 24.915 | 2.584 | 0.146 | 0.030 | 1.398 | 488.7 |
| 2.00 | 0.5 | 24 | 26.458 | 2.687 | 0.146 | 0.022 | 1.461 | 509.5 |
| 3.00 | 0.5 | 25 | 27.128 | 1.854 | 0.144 | 0.030 | 1.358 | 493.8 |
| 3.00 | 1.0 | 8 | 9.456 | 1.807 | 0.112 | 0.028 | 1.423 | 415.6 |
| 0.01 | 3.0 | 1 | 1.012 | 0.109 |  |  |  |  |
| 1.00 | 3.0 | 1 | 1.798 | 0.882 |  |  |  |  |
| 2.00 | 3.0 | 1 | 2.797 | 1.336 |  |  |  |  |
| 3.00 | 3.0 | 1 | 3.751 | 1.635 |  |  |  |  |

The columns are, in order: the Dirichlet Process’s concentration parameter $\alpha$ and the within-cluster variance $\sigma_y^2$ ; the final number of clusters $k$ after assigning each unit to a cluster; the mean number of clusters over the iterations, $mean(k)$ ; the standard deviation of the number of clusters over the iterations, $sd(k)$ ; and the cluster validity measures, Silhouette, Dunn, Davies–Bouldin (DB), and Calinski–Harabasz (CH) indices.

At the end of the algorithm, the cluster memberships over the iterations (1000 iterations) are returned, i.e. the cluster membership distribution. Finally, each observation *i* is assigned to the cluster with the highest probability to represent the cluster membership for that *i*-th observation.

#### Interpretation of the clusters

For each identified cluster, we report the mean of the log MFI-values for each factor (representing a distinct disease), along with the mean age and mean population density.

Additionally, we provide the frequency distribution within each cluster for the following categorical variables: sex (female and male), occupation classes (housewife; none; outside activities, including farming, fishing, and plantation work; students; other), wealth class (Low-income, Lower-middle-income, Middle-income, Wealthy), and vegetation stratum (Dense, Moderate, and Sparse vegetation) [14, 15].

To assess whether the demographic and socio-economic profile of each cluster differed from the rest of the study population, we used Fisher’s exact test for categorical variables, with Monte Carlo-simulated p-values to account for the sparse contingency tables generated by the smaller clusters. For continuous variables, we used the Wilcoxon rank-sum test. All p-values were Bonferroni-corrected.

Finally, for each cluster *k ∈* {1, 2, 3, 4}, we estimated the probability of an observation *i* belonging to that cluster using the following model:

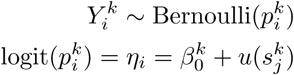

where *Y_i_*is a binary variable assuming value 1 if an observation *i* belongs to cluster *k*, 0 otherwise; 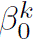 is the global intercept and 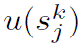 is a Gaussian Random Field (GRF) with a Matérn covariance function [34].

These models were estimated on 1km x 1km prediction grid within the study area (see Figure S3 in the Additional file 1).

## Results

The result of the dimensionality reduction were six factors, each representing a distinct disease. Figure S4 in the Additional file 1 displays the density distribution of the scaled, log-transformed MFI values for the six factors.

Table 2 presents the results of the sensitivity analysis to select the DP concentration parameter *α* and the within-cluster variance 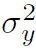, with the optimal values shown in bold. The columns in Table 2 are, in order: the parameter values *α* and 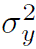; the final number of clusters (*k*) after assigning each unit to a cluster; the mean number of clusters over the iterations (mean(*k*)); the standard deviation of the number of clusters over the iterations (sd(*k*)); and the cluster validity measures (Silhouette, Dunn, DB and CH). Rows where the number of clusters, *k*, equals 1 are left blank, as the internal validation indices cannot be meaningfully computed.

To achieve a more interpretable result, we limited our choice to parameter combinations that produced between 2 and 10 clusters. Among these, the best combination was found to be *α* = 3 and 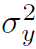 = 1.5, the corresponding number of clusters is *k* = 4 and their numerosities are: Cluster 1 = 5965 observations (96.71%), Cluster 2 = 22 observations (0.36%), Cluster 3 = 157 observations (2.54%), Cluster 4 = 24 observations (0.39%).

Figure 2 shows the average log-MFI values for each disease per cluster and the corresponding numerical values are detailed in Table 3. Seroprevalence estimates and associated risk factors for these same pathogens, based on established antigen-specific cut-offs, have been reported in a previous study [14]. The overall characteristics of the clusters are shown in Figure 3 and in Figure 4 with further details also available in Table 3. Cluster 1 contains the majority of observations, while the other three clusters are considerably smaller.

**Fig 2.**
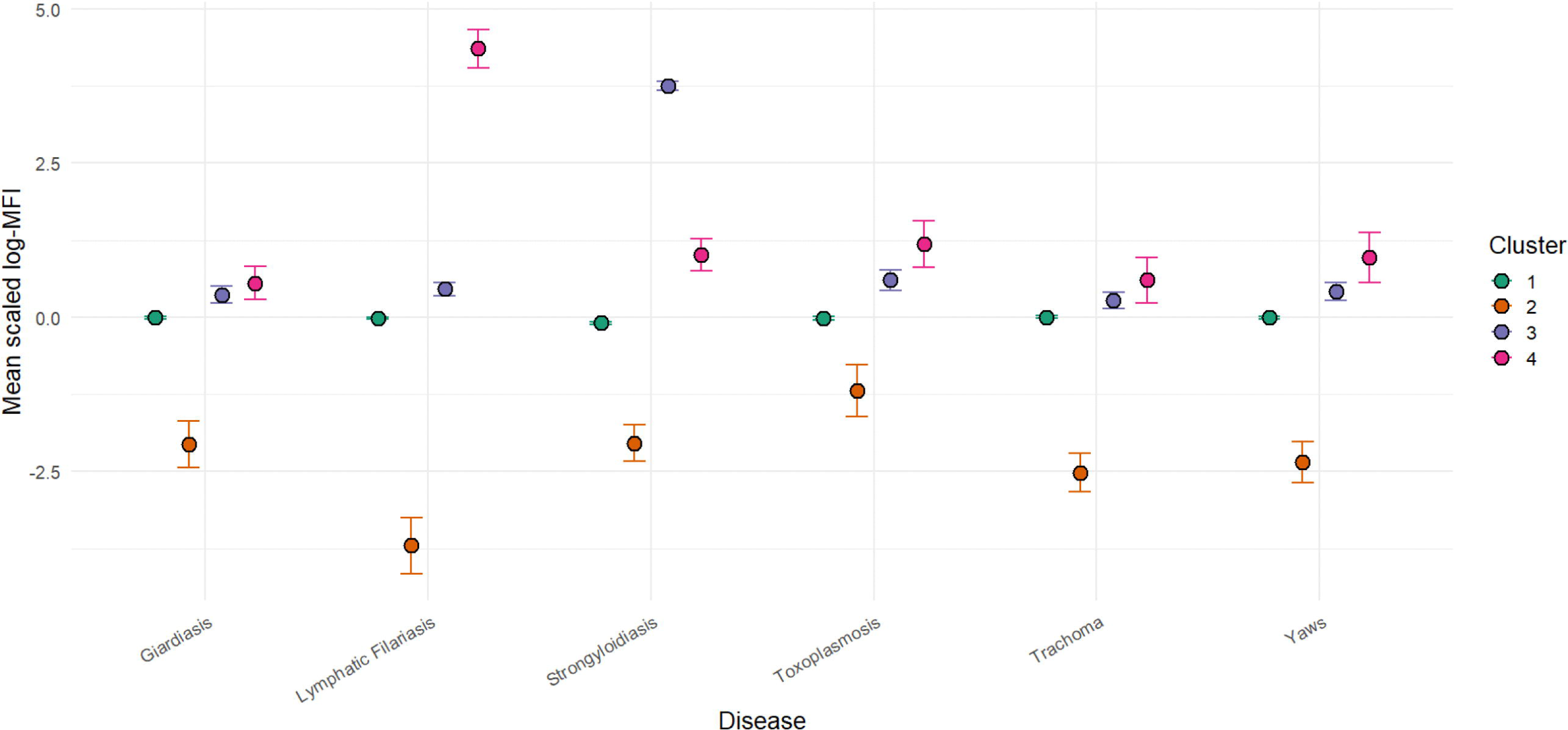
Mean of Disease scaled log-MFI values by Cluster. Lines represent 95% Confidence Intervals.

**Fig 3.**
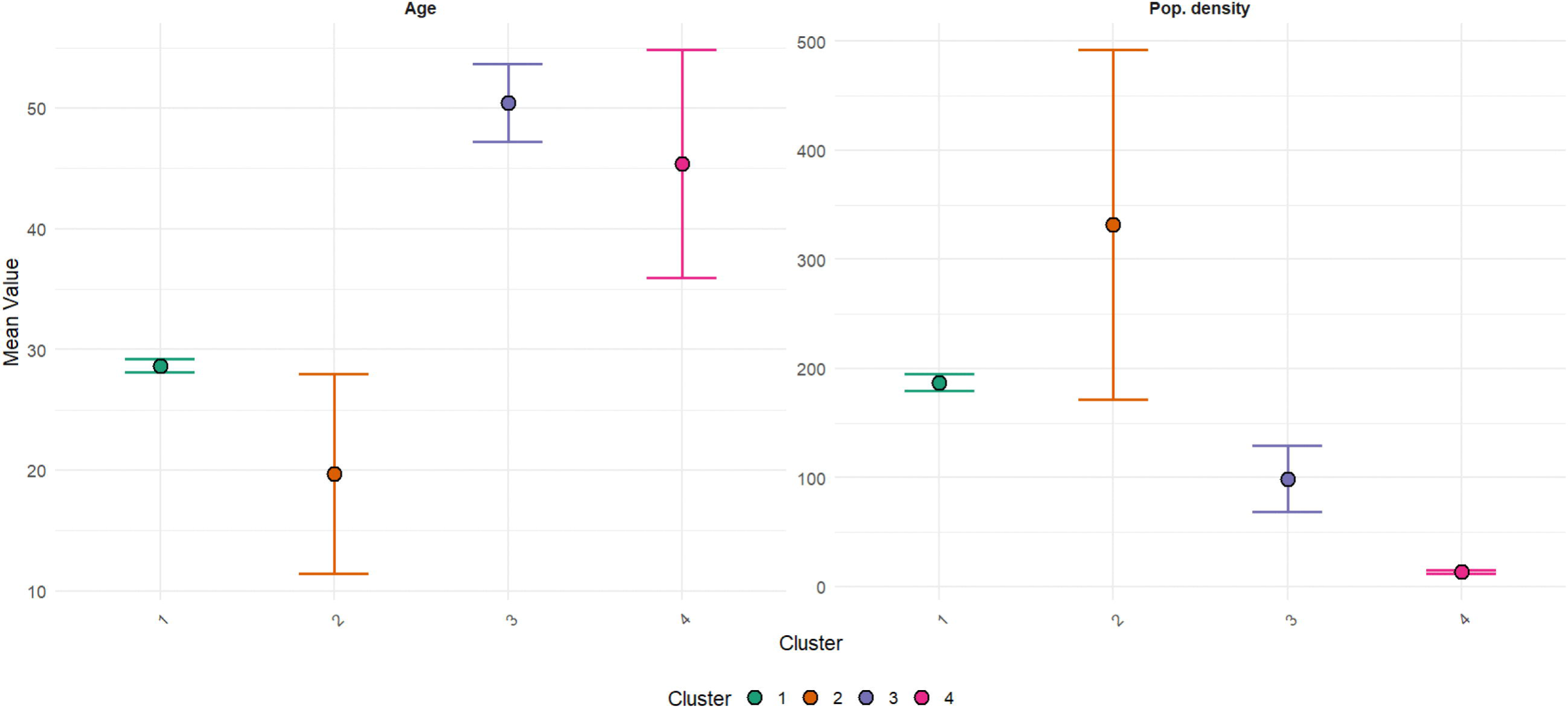
Mean age and mean population density by Cluster. Lines represent 95% Confidence Intervals.

**Fig 4.**
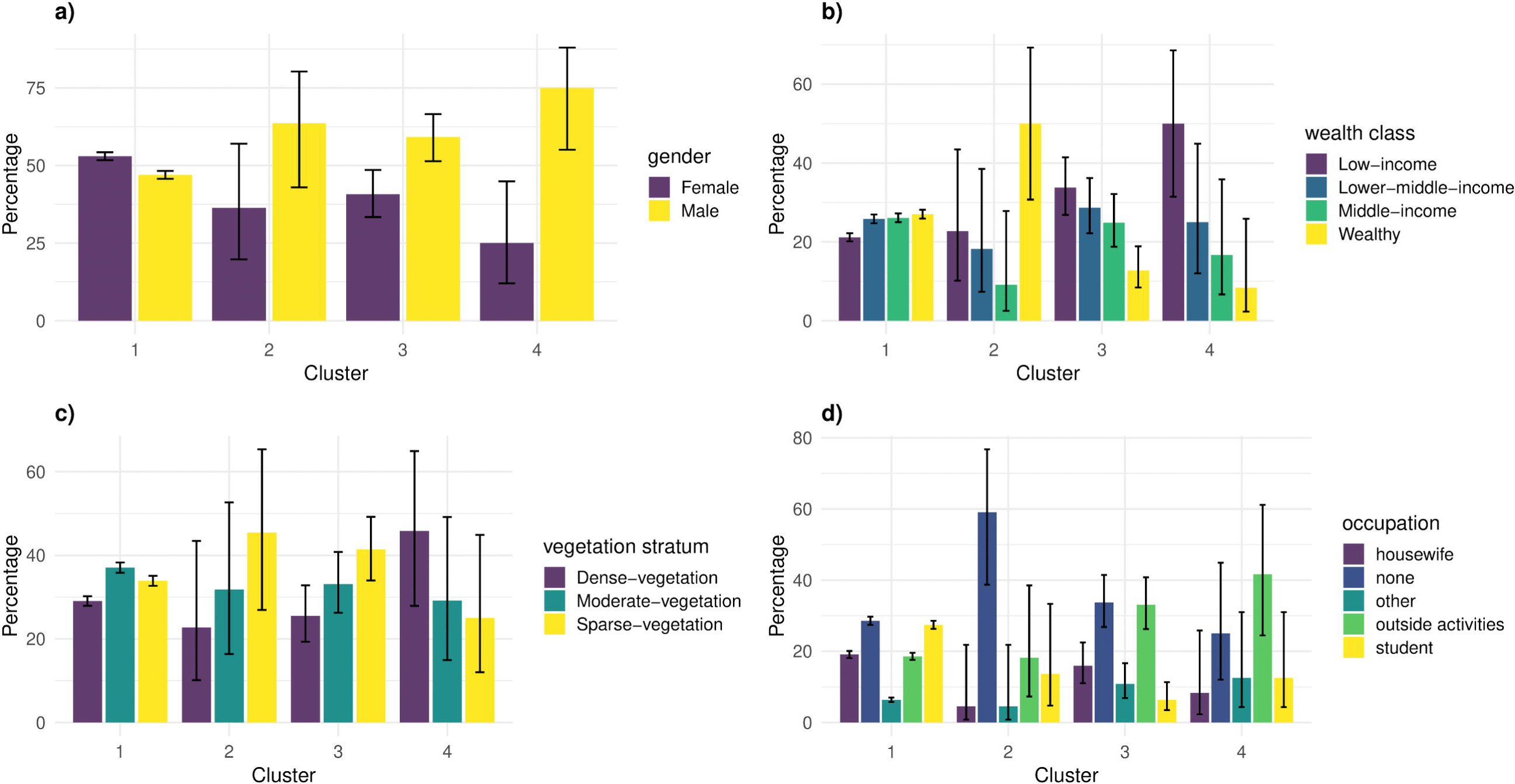
Distribution of gender (a), wealth class (b), vegetation stratum (c) and occupation (d) by Cluster. Lines represent 95% Wilson Confidence Intervals.

**Table 3.** Clusters’ characteristics were presented as the mean and numerosity for continuous variables, and as percentage frequency and numerosity for categorical variables.

| Variable | Cluster 1 |  | Cluster 2 |  | Cluster 3 |  | Cluster 4 |  |
| --- | --- | --- | --- | --- | --- | --- | --- | --- |
|  | N | Mean/Freq | N | Mean/Freq | N | Mean/Freq | N | Mean/Freq |
| **Gender** | 5965 |  | 22 |  | 157* |  | 24* |  |
| Female | 3162 | 53% | 8 | 36% | 64 | 41% | 6 | 25% |
| Male | 2803 | 47% | 14 | 64% | 93 | 59% | 18 | 75% |
| **Age** | 5965 | 29 | 22* | 20 | 157* | 50 | 24* | 45 |
| **Vegetation stratum** | 5965 |  | 22 |  | 157 |  | 24 |  |
| Dense-vegetation | 1733 | 29% | 5 | 23% | 40 | 25% | 11 | 46% |
| Moderate-vegetation | 2210 | 37% | 7 | 32% | 52 | 33% | 7 | 29% |
| Sparse-vegetation | 2022 | 34% | 10 | 45% | 65 | 41% | 6 | 25% |
| **Population Density** | 5965 | 187 | 22 | 332 | 157 | 99 | 24* | 14 |
| **Wealth class** | 5965 |  | 22 |  | 157* |  | 24* |  |
| Low-income | 1260 | 21% | 5 | 23% | 53 | 34% | 12 | 50% |
| Lower-middle-income | 1539 | 26% | 4 | 18% | 45 | 29% | 6 | 25% |
| Middle-income | 1555 | 26% | 2 | 9% | 39 | 25% | 4 | 17% |
| Wealthy | 1611 | 27% | 11 | 50% | 20 | 13% | 2 | 8% |
| **Occupation** | 5965 |  | 22 |  | 157* |  | 24 |  |
| Housewife | 1138 | 19% | 1 | 5% | 25 | 16% | 2 | 8% |
| None | 1704 | 29% | 13 | 59% | 53 | 34% | 6 | 25% |
| Other | 380 | 6% | 1 | 5% | 17 | 11% | 3 | 12% |
| Outside activities | 1107 | 19% | 4 | 18% | 52 | 33% | 10 | 42% |
| Student | 1636 | 27% | 3 | 14% | 10 | 6% | 3 | 12% |
| **Toxoplasmosis** | 5965 | -0.016 | 22 | -1.2 | 157 | 0.6 | 24 | 1.2 |
| **Giardiasis** | 5965 | -0.0042 | 22 | -2.1 | 157 | 0.37 | 24 | 0.55 |
| **Lymphatic Filariasis** | 5965 | -0.016 | 22 | -3.7 | 157 | 0.46 | 24 | 4.4 |
| **Trachoma** | 5965 | -0.00031 | 22 | -2.5 | 157 | 0.27 | 24 | 0.6 |
| **Strongyloidiasis** | 5965 | -0.095 | 22 | -2 | 157 | 3.8 | 24 | 1 |
| **Yaws** | 5965 | -0.0062 | 22 | -2.3 | 157 | 0.42 | 24 | 0.97 |

\* indicates a statistically significant difference ( $p < 0.05$ , Bonferroni-corrected) between the given cluster and the rest of the study population. Categorical variables were tested using Fisher’s exact test (Monte Carlo-simulated, 10,000 replicates); continuous variables were tested using the Wilcoxon rank-sum test.

As illustrated in Figure 2, Cluster 2 is distinguished by lower mean values across all diseases compared to the other clusters. Conversely, Clusters 3 and 4 are characterized by a higher mean value of strongyloidiasis and lymphatic filariasis, respectively.

As shown in Figure 3, people in Cluster 2 have a lower mean age and a higher population density compared to the other clusters. Additionally, this cluster also has a higher percentage of wealthy individuals (Figure 4, panel b), and a greater proportion of people residing in areas with sparse vegetation (Figure 4, panel c). Cluster 2 also contains a higher percentage of people with no occupation compared to other occupation types (Figure 4, panel d).

In contrast to the trends observed in Cluster 2, Clusters 3 and 4 show an opposite pattern in key characteristics. As shown in Figure 3, these clusters have a higher mean age and a lower population density value compared to the other clusters. A look at Figure 4, panel b, reveals that Clusters 3 and 4 have a higher percentage of individuals in the low-income and lower-middle-income classes. These clusters also contain a greater proportion of people who perform outdoor activities such as farming, fishing, and working in plantations (Figure 4, panel d). Additionally, Cluster 4 is characterized by a higher percentage of people living in dense vegetation areas (Figure 4, panel c).

There is no significant difference between the percentage of males and females in most clusters, with the exception of Cluster 3 and 4, which is characterized by a higher percentage of men (Figure 4, panel a).

Cluster 1 has no distinct characteristics, as its mean age and population density are similar to the overall sample’s, and there are no significant differences in the frequency distribution across its categorical variables.

Formal testing (Table 3) showed that Cluster 2 differed significantly from the rest of the sample only in age (*p* = 0.043); Cluster 3 differed significantly in gender (*p* = 0.010), wealth class (*p <* 0.001), occupation (*p <* 0.001) and age (*p <* 0.001) distribution.

Cluster 4 differed significantly in gender (*p* = 0.022), wealth class (*p* = 0.025), age (*p* = 0.002) and population density (*p <* 0.001).

Figure 5 shows the spatial distribution of the predicted probability of a household belonging to each cluster within the study area. Cluster 2 didn’t display a clear spatial distribution, likely due to its small size; location of households belonging to cluster 2 is detailed in Figure S5 of the supplementary materials.

**Fig 5.**
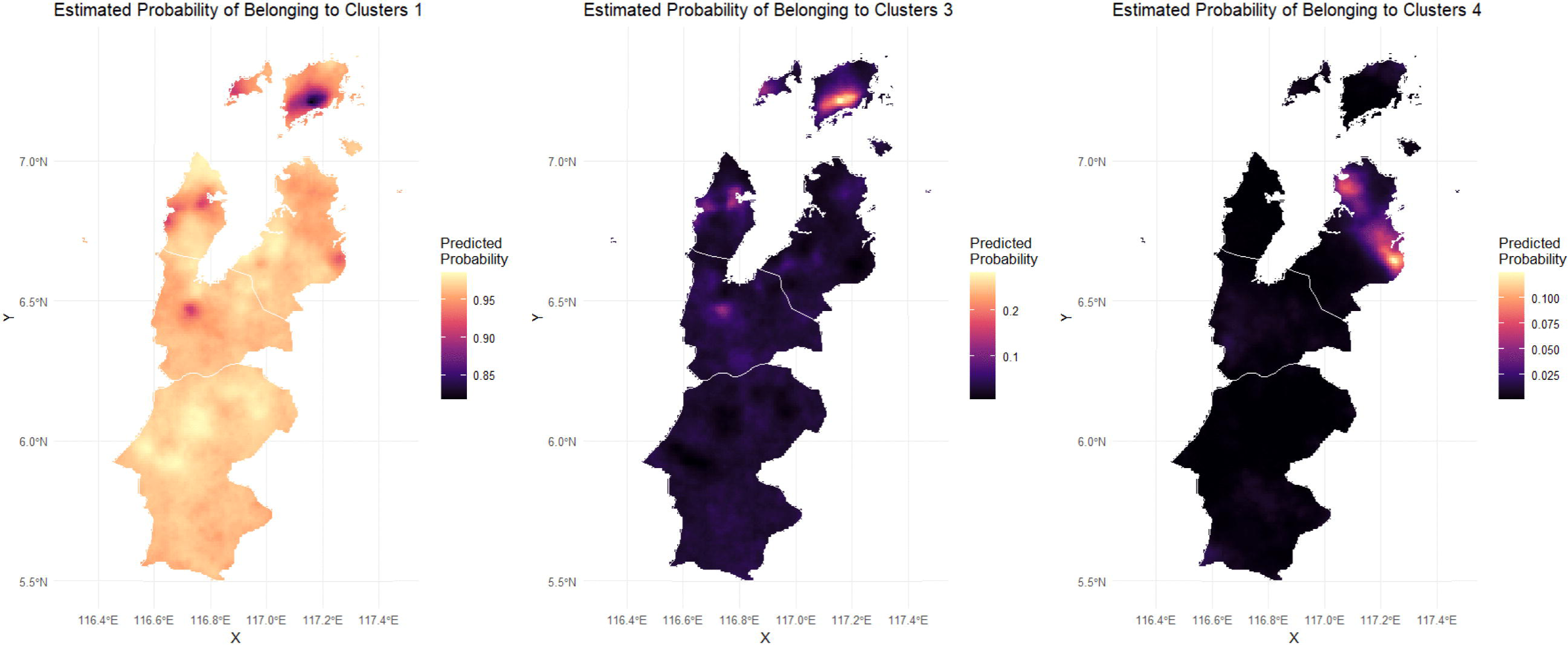
Estimated probability of belonging to each cluster.

Looking at Figure 5, there’s a very high probability of belonging to Cluster 1 across the entire study area. This finding reflects the large size of this cluster. The only exceptions are certain darker areas in the map for Cluster 1, which correspond to the brightest spots in the maps for Cluster 3 and Cluster 4. These bright areas indicate locations with a higher predicted probability of a household belonging to Cluster 3 or 4.

## Discussion

This study applies model-based cluster analysis to quantitative multiplex serology data, enabling the simultaneous analysis of antibody responses to all pathogens included in the assay. To the best of our knowledge, this is the first study to use this approach to identify coherent cross-pathogen exposure clusters. The analysis reveals distinct sub-populations characterised by consistent multi-pathogen profiles, alongside clear socioeconomic and spatial patterns.

Model-based clustering provides a principled statistical framework for uncovering latent sub-groups within the population by assuming that the observed data arise from a mixture of underlying probability distributions. In our setting, this allows the algorithm to identify groups of individuals who share similar antibody response profiles across all pathogens measured in the multiplex assay. Importantly, the clusters are learned directly from the joint distribution of the data, without prespecifying relationships among pathogens or adjusting for factors such as age or socioeconomic status in advance.

In the application presented in this work the algorithm detected four sub-populations, whose characteristics showed strong consistency with the serological profiles and align well with the epidemiological literature: individuals in Cluster 2 exhibited lower mean quantitative antibody levels across all measured diseases compared to other groups.

They tended to reside in high-population density areas, possibly corresponding to larger villages with potentially better living and sanitary conditions, which could explain the generally lower pathogen exposure. Furthermore, this cluster was on average younger, reflecting the established relationship between age and cumulative exposure to NTDs infections, where older age typically correlates with higher seroprevalence [22].

On the other hand, Clusters 3 and 4 were characterized by significantly higher mean MFI values for specific helminths: strongyloidiasis (Cluster 3) and lymphatic filariasis (Cluster 4). Individuals in both clusters were on average older and tended to reside in low-population density areas, potentially indicative of poorer living conditions. Crucially, people in these groups reported primarily engaging in outdoor activities such as farming, fishing, and plantation work. Such occupations are known risk factors, linking direct contact with infected soil to strongyloidiasis [24, 35] and exposure to mosquitoes to lymphatic filariasis [36]. Cluster 4, specifically, showed a higher percentage of people living in dense vegetation areas, which is a recognized environmental risk factor for lymphatic filariasis [37]. The estimated spatial model provides a way to identify geographic areas where households belonging to each cluster are concentrated. The spatial distribution of the estimated probability maps for Clusters 3 corresponds to the areas of high mean posterior estimated seroprevalence for lymphatic filariasis previously reported by Chan et al. [14] using the same data. The consistency of our model results with existing literature validates its potential as a tool for informing area-specific interventions and risk monitoring. The maps are inherently complementary because cluster membership is mutually exclusive.

The distinct profiles identified could be used to develop actionable public health strategies that target multiple co-occurring NTDs simultaneously. Cluster 4 shows its highest exposure to lymphatic filariasis, alongside elevated antibody levels for giardiasis, strongyloidiasis, toxoplasmosis, trachoma, and yaws relative to the rest of the population, and is characterised by residence in densely vegetated areas; here, LF-specific mass drug administration could be delivered alongside treatment for these other NTDs in a single integrated visit, and could be further complemented by environmental vector control measures targeting mosquito breeding sites in the surrounding area. Cluster 3 shows its highest exposure to strongyloidiasis, together with elevated levels of the remaining five diseases, and is characterised by older age and outdoor occupations such as farming and fishing; here, the priority intervention could instead centre on ivermectin-based mass treatment and improved sanitation infrastructure in agricultural and fishing areas, again bundled with treatment for the other co-elevated pathogens rather than delivered as a standalone STH campaign. Cluster 2, by contrast, shows consistently lower exposure across all six pathogens and a younger, wealthier profile, suggesting these areas could be assigned lower priority, allowing integrated intervention resources to be concentrated in Clusters 3 and 4. Cluster 1, comprising the large majority of the study population, shows no distinctive serological, demographic, or socioeconomic profile. While it does not point to a specific intervention, it serves as the baseline against which the elevated-risk profiles of Clusters 2, 3, and 4 are defined, and its relative size could be monitored over time as a simple indicator of programme impact. As targeted interventions reduce residual transmission in Clusters 3 and 4, the proportion of the population with an undifferentiated, low-risk profile would be expected to grow, thus making Cluster 1 bigger in size.

Several limitations of our study should be carefully considered. As with any analysis of quantitative serological data, the proposed approach is influenced by assay characteristics, including limits of detection and dynamic range. However, because the analysis targets relative response patterns across multiple antigens rather than absolute thresholds for individual markers, the clustering is primarily driven by shared antibody profiles rather than assay-specific scaling. In this context, technical effects are more likely to affect the overall dispersion of the data than the underlying cluster structure. Nevertheless, appropriate normalisation and basic sensitivity analyses remain essential components of the presented analytical workflow.

The analysis of the spatial distribution of clusters was presented here as a descriptive approach to aid interpretation of cluster distribution across the study area, rather than as a fully calibrated spatial inferential model. We modelled the probability of belonging to each cluster using four independent Bernoulli models, each including a Gaussian Random Field. A more fundamental limitation of this approach is that cluster membership itself is estimated with uncertainty under the DPMM, rather than observed as a fixed label. A fully coherent extension of this framework would therefore need to embed the spatial random field directly within the clustering process, for instance through spatially dependent formulations of the Dirichlet process, so that cluster allocation and spatial structure are estimated jointly, with uncertainty propagated consistently between the two stages. Developing and validating such an approach represents a substantial methodological undertaking and an important direction for future work.

Our selection of the DPMM parameters *α* and *σ*^2^ was guided by a combination of internal validity indices and interpretability, following the approach used by [33]. Rather than choosing the combination that optimised any single index, we restricted our search to configurations yielding between 2 and 10 clusters, since larger numbers of clusters produced groups too small to characterise meaningfully. Among these, the chosen combination (*α* = 3, *σ*^2^ = 1.5) performed best across most validity indices among the *k* = 4 solutions, and, critically, resolved distinct disease-specific sub-populations consistent with previously reported spatial patterns of exposure in this population [14], unlike the alternative *k* = 2 solution. This mirrors the logic of [33], who similarly found that configurations optimising a single index in isolation could fragment a true cluster into spurious sub-groups, and recommended that substantive interpretability guide parameter selection alongside quantitative criteria.

As in any serological data analysis, age is an important factor to consider as it is directly linked to cumulative antibody levels and may strongly influence the serological profiles. To assess this, we applied the methods described above across three distinct age classes: 0-15 years old (2288 people), 16-30 years old (1186 people) and over 30 years old (2694 people). As demonstrated in the Supplementary Materials (see Additional file 1, Tables S2–S4 and Figures S6–S8), the core serological and socioeconomic patterns remained largely consistent, suggesting the patterns are not solely driven by age stratification. However, the interpretation of age-dependent variables, such as occupation, was naturally influenced by this classification. Moreover, the frequency distribution of variables associated with clusters containing fewer than 20 people were difficult to interpret, as the resulting percentages may reflect data noise rather than true sample characteristics.

A further limitation is connected to the interpretation of Cluster 1: because it is the largest cluster, encompassing the majority of observations, it lacks the distinct, specific characterization observed in the smaller clusters. Future work should therefore focus on developing methodologies to help disaggregate and characterize the internal heterogeneity of such large, non-specific clusters.

Finally, the literature offers an alternative approach to selecting the Dirichlet Process concentration parameter *α*, treating it as a random hyperparameter with its own prior distribution and estimating it jointly with the mixture components within the Gibbs sampler, via a data augmentation step [38]. This differs fundamentally from the approach adopted here, in which *α* and *σ*^2^ are treated as fixed hyperparameters, selected externally through a sensitivity analysis over a grid of candidate values compared using internal validity indices. A fully Bayesian treatment of *α* could make the use of DPMM more efficient, but is unlikely to reduce the subjectivity involved in this modelling step, since it introduces its own prior specification choices and additional computational complexity, without necessarily guaranteeing improved clustering performance in every application. The sensitivity analysis used in this work, by contrast, offers a comparatively simple and transparent procedure, requiring no additional prior elicitation or changes to the sampling algorithm, and remains straightforward for practitioners to implement and interpret. Ultimately, fully replicated analyses in diverse epidemiological settings will be required to support the wider applicability of the chosen or alternative approaches to inform the selection of *α* and *σ*^2^.

While acknowledging current limitations, this study serves as a successful proof-of-concept for applying model-based clustering to multiplex serology data: the method was able to detect consistent and epidemiologically plausible patterns linking quantitative antibody levels, geographical position, and socioeconomic characteristics. Beyond the methodological contribution, these findings offer a practical framework for monitoring infectious disease risk and for informing programmatic approaches to NTD elimination. Rather than applying uniform, district-wide interventions, our approach can help identify specific geographic areas and population subgroups that bear an elevated disease burden, supporting more targeted strategies, particularly in the post-MDA surveillance phase when residual transmission is often confined to localised hotspots. Because our method jointly characterises multiple pathogens, it can also enhance integrated surveillance platforms by guiding more tailored, multi-disease sample collection and treatment protocols.

## Supporting information

**Additional file 1 Table S1.** Descriptive statistics of the socio-economics variables.

**Figure S1**. Correlation matrix antigens’ scaled log-MFI values.

**Figure S2.** Scatterplot between the study variables.

**Figure S3.** 1 km × 1 km prediction grid within study area.

**Figure S4.** Scaled-log MFI distribution by Diseases.

**Table S2.** Clusters’ characteristics.

**Figure S5.** Location of households belonging to cluster 2.

**Table S3.** Resulting parameters from the sensitivity analysis and number of observations per cluster in age class 0-15 years old.

**Figure S6.** Mean of Disease and socio-economic characteristics by cluster in age class 0-15 years old.

**Table S4.** Resulting parameters from the sensitivity analysis and number of observations per cluster in age class 16-30 years old.

**Figure S7.** Mean of Disease and socio-economic characteristics by cluster in age class 16-30 years old.

**Table S5.** Resulting parameters from the sensitivity analysis and number of observations per cluster in age class over 30 years old.

**Figure S8.** Mean of Disease and socio-economic characteristics by cluster in age class over 30 years old.

## Abbreviations

MBA: Multiplex Beads Assays
IgG: Immunoglobulin G
PCA: Principal Component Analysis
LF: Lymphatic Filariasis
NTDs: Neglected Tropical Diseases
MFI-bg: Median fluorescence intensity minus background
DPMM: Dirichlet Process mixture model
DP: Dirichlet Process
DB: Davies–Bouldin index
CH: Calinski–Harabasz index
GRF: Gaussian Random Field

## Acknowledgments

We would like to thank the Director-General, Ministry of Health, Malaysia, for permission to publish this manuscript. We also thank the members of the Task Force for Global Health, Dr. Katie Gass, Dr. Kumar Udhayakumar, Dr. Patrick Lammie and Dr. Kimberly Fornace, for their expert review and valuable contributions to the quality of this work.

## Ethics statement

The Medical Research Sub-Committee of the Malaysian Ministry of Health (NMRR-14-713-21117) and the Research Ethics Committee of the London School of Hygiene and Tropical Medicine (8340) approved the Malaysian study and written informed consent was obtained from all participants. For minors, written informed consent to participate in this study was provided by the participants’ legal guardian/next of kin, as described in previous works [14, 15]. Because the authors did not interact with study participants and had no access to personally identifiable information, they were determined to be ‘not engaged’ in human subjects research.

## Data availability

Simulated data and scripts allowing the reproducibility of the methodology presented in this study are publicly available on GitHub. Please note that because the repository relies on simulated data created to mirror the structure of the original dataset, running these analyses will yield numerical results and plots that are not identical to those in the published paper.

