## Additional file 1 for "Modelling serological multiplex bead assays responses: A case study from Malaysia"

### i Data description

Table S1: Descriptive statistics of the socio-economics variables

| Variable | N | Mean/% | Std. Dev. | Median | Min | IQR | Max |
|---|---|---|---|---|---|---|---|
| **Sex** | 6168 | | | | | | |
| Female | 3240 | 53% | | | | | |
| Male | 2928 | 47% | | | | | |
| **Vegetation stratum** | 6168 | | | | | | |
| Dense vegetation | 1789 | 29% | | | | | |
| Moderate vegetation | 2276 | 37% | | | | | |
| Sparse vegetation | 2103 | 34% | | | | | |
| **Wealth class** | 6168 | | | | | | |
| Low-income | 1330 | 22% | | | | | |
| Lower-middle-income | 1594 | 26% | | | | | |
| Middle-income | 1600 | 26% | | | | | |
| Wealthy | 1644 | 27% | | | | | |
| **Occupation** | 6168 | | | | | | |
| housewife | 1166 | 19% | | | | | |
| none | 1776 | 29% | | | | | |
| other | 401 | 7% | | | | | |
| outside activities | 1173 | 19% | | | | | |
| student | 1652 | 27% | | | | | |
| **Age** | 6168 | 29 | 22 | 25 | 0.22 | 35 | 105 |
| **Population density** | 6168 | 185 | 298 | 34 | 0 | 190 | 1259 |

### ii Dimensionality reduction

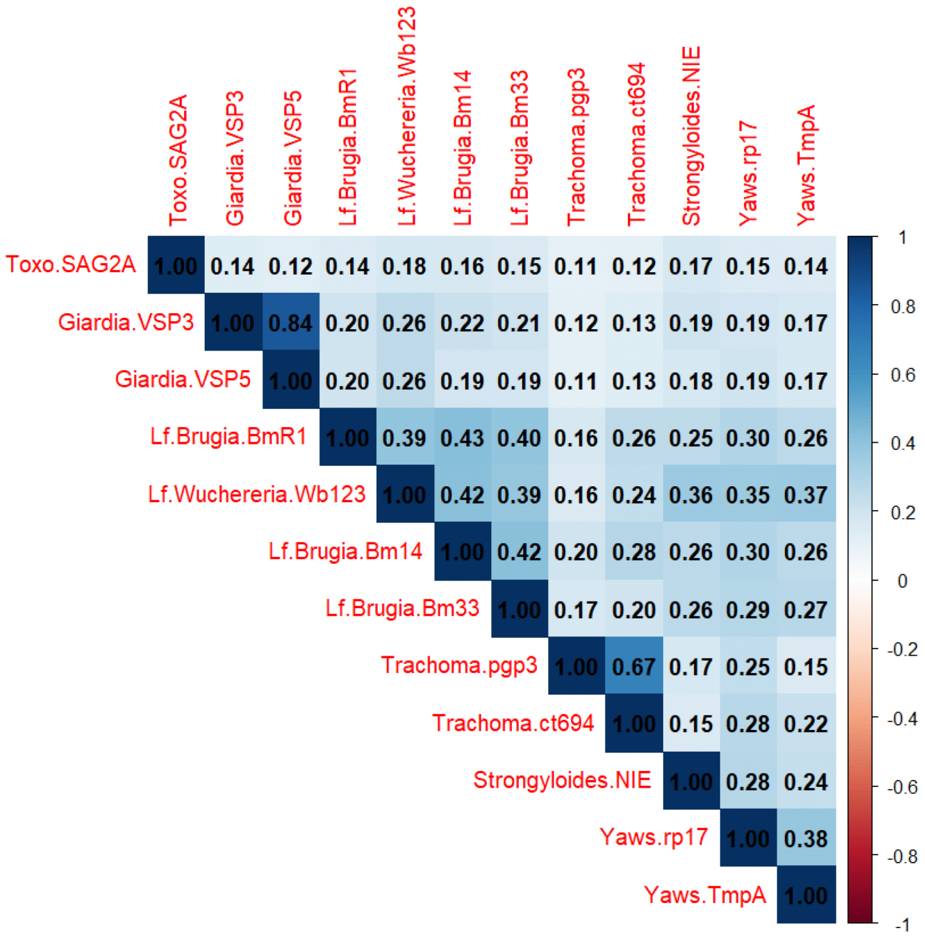


Figure S1: Correlation matrix antigens' scaled log-MFI values.

### iii Cluster analysis

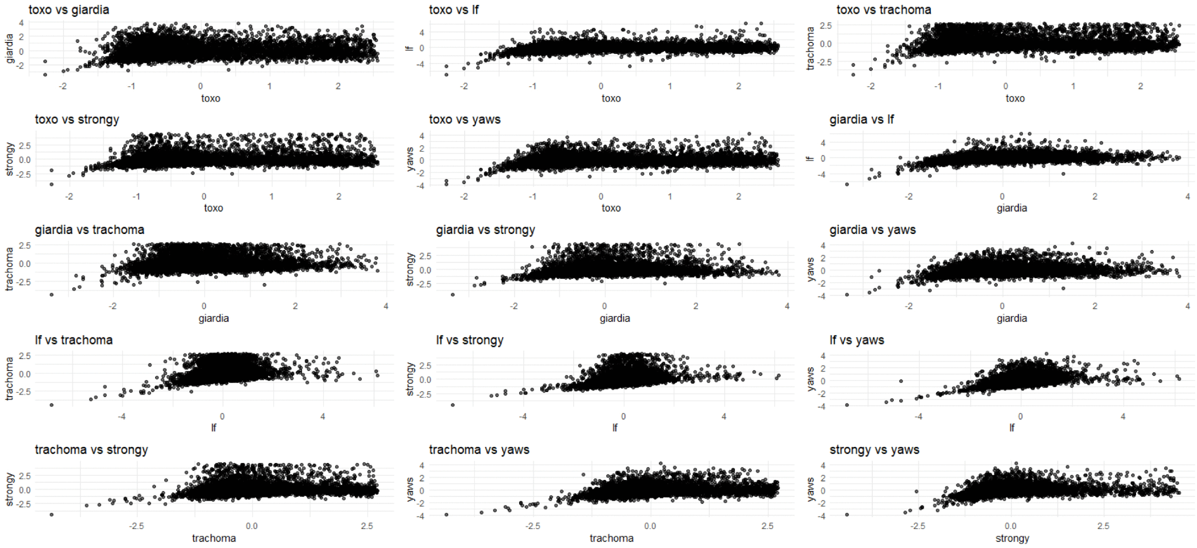


Figure S2: Scatterplot between the study variables.

### iv Geospatial modelling

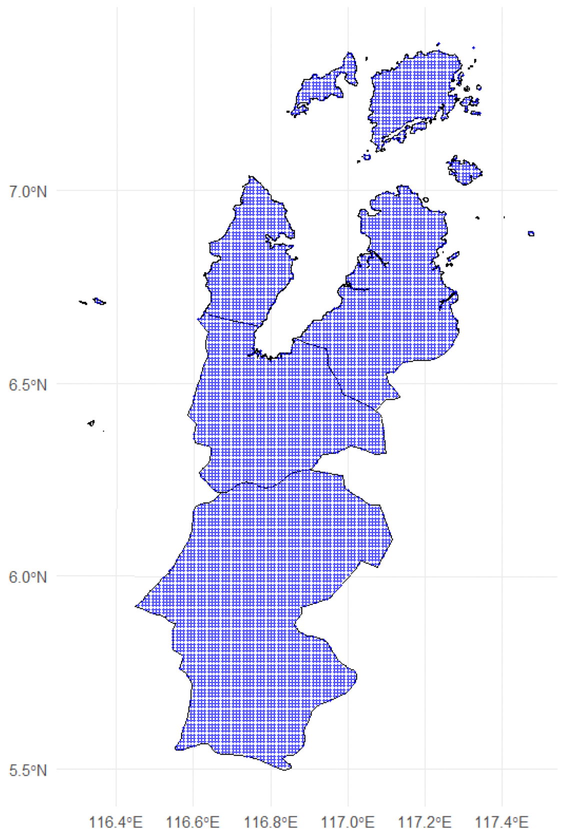


Figure S3: 1 km × 1 km prediction grid within study area, Sabah (Malaysia).

### v Results

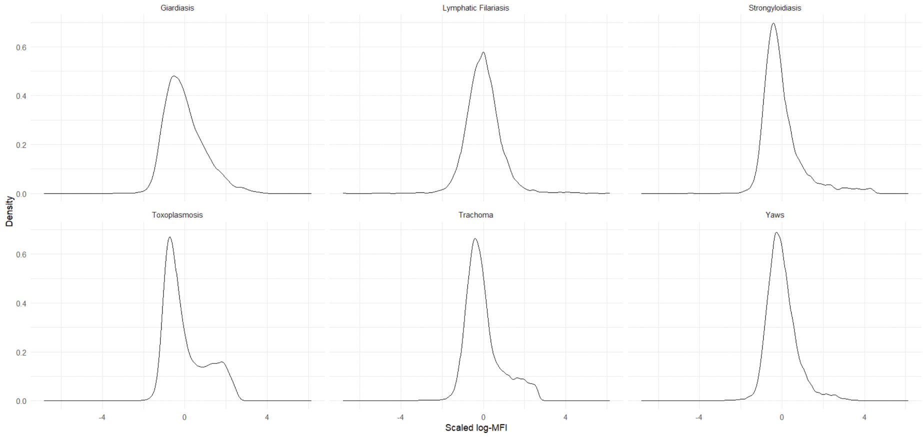


Figure S4: Scaled-log MFI distribution by Diseases.

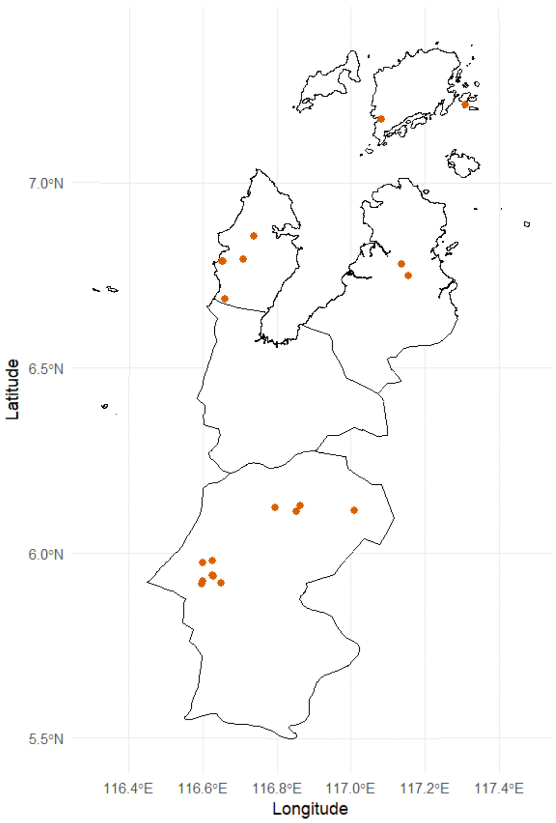


Figure S5: Location of households belonging to cluster 2 over the study area, Sabah (Malaysia).

### vi Methodology applied to age classes

The age brackets (0–15, 16–30, and > 30 years) were selected to capture distinct epidemiological stages: the 0–15 age group serves as a standard benchmark for assessing recent ongoing transmission dynamics and the impact of recent control interventions, whereas the older age classes reflect progressive levels of historical cumulative exposure in young and older adults.

| $\alpha$ | $\sigma_y^2$ | $n_1$ | $n_2$ |
|---|---|---|---|
| 0.01 | 2 | 2279 | 9 |

Table S2: On the left: Resulting $\alpha$ and $\sigma_y^2$ from the sensitivity analysis. On the right: Number of observations per cluster, age class 0-15 years old.

| $\alpha$ | $\sigma_y^2$ | $n_1$ | $n_2$ |
|---|---|---|---|
| 1 | 1.5 | 1180 | 6 |

Table S3: On the left: Resulting $\alpha$ and $\sigma_y^2$ from the sensitivity analysis. On the right: Number of observations per cluster, age class 16-30 years old.

| $\alpha$ | $\sigma_y^2$ | $n_1$ | $n_2$ | $n_3$ | $n_4$ |
|---|---|---|---|---|---|
| 1 | 1.5 | 2559 | 10 | 119 | 6 |

Table S4: On the left: Resulting $\alpha$ and $\sigma_y^2$ from the sensitivity analysis. On the right: Number of observations per cluster, age class over 30 years old.

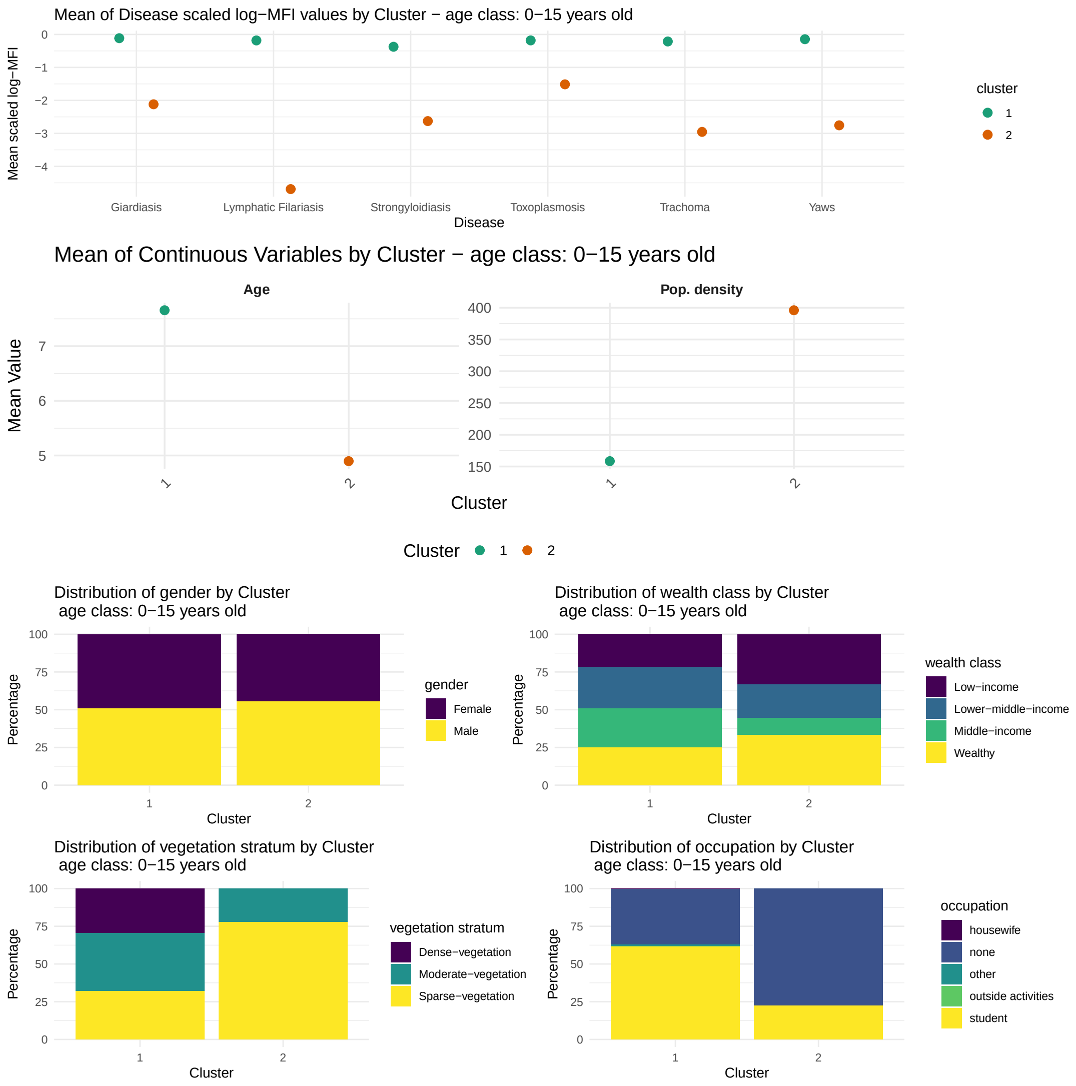


Figure S6: Mean of Disease and socio-economic characteristics by Cluster, age class 0-15 years old.

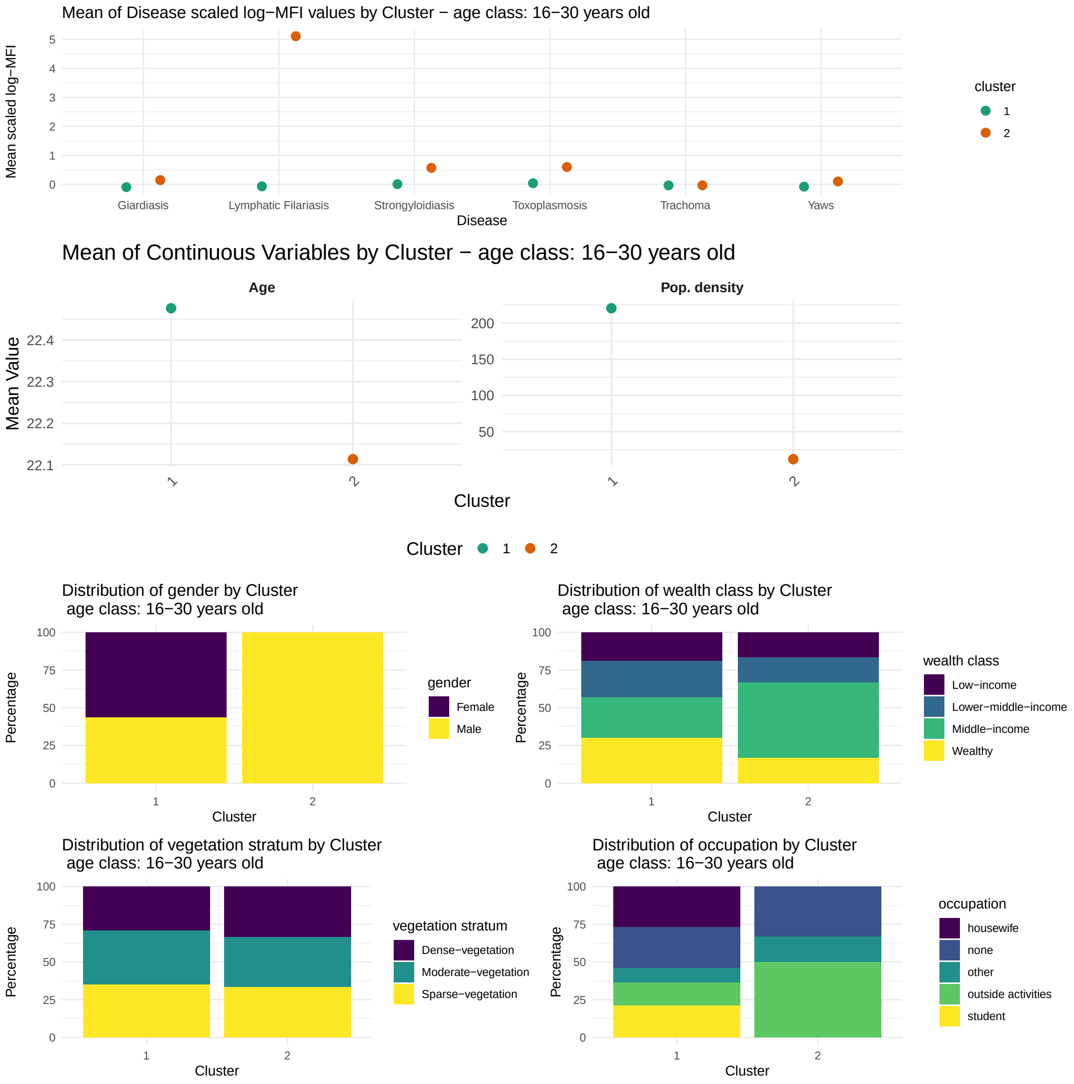


Figure S7: Mean of Disease and socio-economic characteristics by Cluster, age class 16-30 years old.

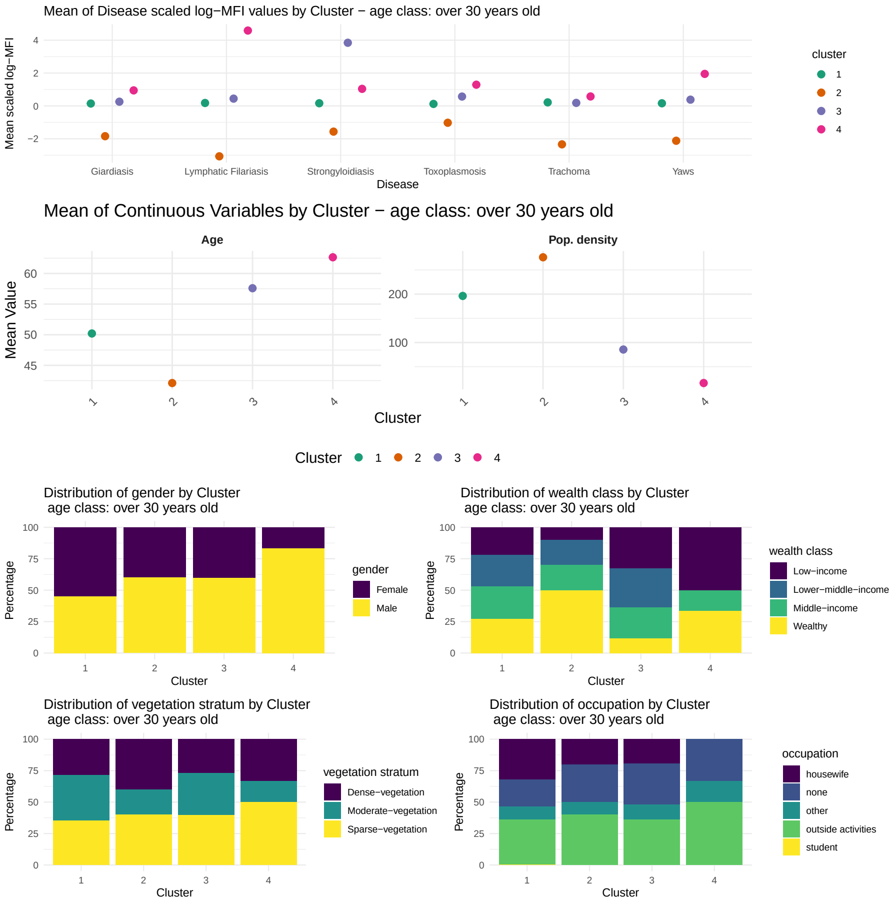


Figure S8: Mean of Disease and socio-economic characteristics by Cluster, age class over 30 years old.
